# Characterising school-age health and function in rural Zimbabwe using the SAHARAN toolbox

**DOI:** 10.1101/2021.09.22.21263533

**Authors:** Joe D Piper, Clever Mazhanga, Gloria Mapako, Idah Mapurisa, Tsitsi Mashedze, Eunice Munyama, Marian Mwapaura, Dzivaidzo Chidhanguro, Eddington Mpofu, Batsirai Mutasa, Melissa J Gladstone, Jonathan C Wells, Lisa F Langhaug, Naume V Tavengwa, Robert Ntozini, Andrew J Prendergast

## Abstract

**Introduction:** School-age health, growth and development are poorly characterised in low- and middle-income countries. We developed the School-Age Health, Activity, Resilience, Anthropometry and Neurocognitive (SAHARAN) toolbox to measure growth, cognitive and physical function in rural Zimbabwe.

**Methods:** The SAHARAN toolbox was developed using a stepwise approach, with tool selection based on COSMIN principles. Growth was measured with anthropometry, skinfold thicknesses and bioimpedance analysis to obtain lean mass index (LMI) and phase angle. Cognition was assessed using a fine motor finger tapping task, school achievement test (SAT), and the Kaufmann Assessment Battery for Children (KABC-2) to determine the mental processing index (MPI). Physical function combined grip strength, broad jump and the 20m shuttle run test to produce a total physical score (TPS). A detailed caregiver questionnaire was performed in parallel.

**Results:** 80 Zimbabwean children with mean (SD) age 7.6 (0.2) years had mean height-for-age (HAZ) and weight-for-age Z-scores (WAZ) of −0.63 (0.81) and −0.55 (0.85), respectively. For growth measures, LMI and total skinfold thicknesses were highly related to both WAZ and BMI Z-score, but not to HAZ. For physical function, TPS was associated with unit rises in HAZ (1.29, 95%CI 0.75,1.82, p<0.001), and LMI (0.50, 95%CI 0.16,0.83, p=0.004), but not skinfold thicknesses. For cognition, the SAT was highly associated with unit increases in the MPI (0.9 marks, 95%CI 0.4,1.3, p<0.001) and a child socioemotional questionnaire (8.2 marks, 95%CI 2.9,13.5, p=0.003). Phase angle was associated with 3.5 seconds quicker time to complete the finger tapping task (95%CI 0.6,6.4, p=0.02). No child outcomes were associated with socioeconomic status, nurturing, discipline, food and water insecurity, or household adversity.

**Conclusions:** We found clear associations between growth, height-adjusted lean mass and physical function, but not cognitive function, in a cohort of Zimbabwean children. The SAHARAN toolbox could be deployed to characterise school-age growth, development and function in sub-Saharan Africa.

**What is already known?:**

- There are a lack of available tools to measure school-age health, growth, physical function and cognitive function together, particularly in low-resource settings.
- Early-life quality of growth is associated with future chronic disease risk.

**What are the new findings?:**

- School-age markers of growth and lean mass (but not fat mass) were strongly associated with physical function.
- Cognitive function was related to schooling and to the child’s perceived socioemotional score.

**What do the new findings imply?:**

- The SAHARAN toolbox enables an integrated assessment of school-age health, growth, physical and cognitive function.
- The quality of early-life growth and relative lean mass is important at school-age, since it is associated with better physical function and reduced chronic disease risk.

## INTRODUCTION

There is an urgent need to reposition child health within the “survive, thrive and transform” global strategy^1^. This calls for a more holistic approach, recognising all stages of childhood as important for growth and development within a ‘life-course’ perspective. This framework recognises the negative exposures and positive opportunities that occur from conception to adulthood, with each life-stage building on previous stages^2^. It also highlights the interconnectedness and differences between growth, health, physical and cognitive function, allowing interventions to target multiple areas of development.

There has been remarkably little focus on school-age health outcomes between 5-14 years, particularly in low- and middle-income countries (LMICs), since routine health information systems do not capture information in this age group^3^. Mortality at age 5-14 years is disproportionately concentrated in LMICs, particularly sub-Saharan Africa^4^. Beyond survival, less is known about child growth and developmental trajectories after 5 years compared to younger ages, although recent studies have suggested that trends in height and BMI are highly variable in response to different social, nutritional and environmental factors, and are predictive of future health^5 6 7-9^. In particular, a recent study comparing global BMI trends showed that healthy school-age growth can either consolidate gains from early childhood or mitigate nutritional imbalances^9^. Conversely, in some countries (e.g. South Africa), children exhibit negative trajectories in health with a relative decline in height and an increase in BMI^9^. There is a pressing need to better understand the impact of school-age risk factors and protective factors, and the ability to mitigate early disadvantages to improve growth and development trajectories.

Refocusing attention on school-age health outcomes would increase our understanding of the timing of effective interventions to address growth, physical, cognitive, and socioemotional development in LMICs^10^. Following early-life nutrition interventions, some improvements in cardiovascular risk factors^11 12^, cognition^13^ and later human capital^14^ have been reported. However, growth itself is a poor proxy for cognitive function^13^. Few studies have confirmed whether early growth gains translate into long-term improvements in both cognitive and physical function and, conversely, whether interventions showing little or no effect at early ages may nevertheless confer meaningful benefits on later measures of growth and function. For example, in the INCAP trial, children receiving nutritional supplementation into early childhood had higher adult IQ scores, greater work capacity and earnings (among men) and greater schooling (among women)^14^. At school-age, it becomes easier to undertake more sensitive and specific assessments of cognitive development, compared to younger ages. Additionally, certain domains of brain function only begin to develop from 2 years of age, including expressive language and higher cognitive functions such as socio-emotional behaviour. To evaluate the effects of improved nutrition on all aspects of neurodevelopment therefore requires assessment at older ages. For example, follow-up of a trial of lipid-based nutritional supplements (LNS) delivered between 6-18 months in Ghana showed an effect on socio-emotional development at 4-6 years of age^15^, which would not be measurable at 2 years. School-age assessments are more predictive of adult IQ and cognitive function than are early-life assessments, particularly as executive function can be better assessed^16^. Finally, school performance is itself a valuable outcome that can only be evaluated at older ages.

One reason for limited school-age data is a shortage of feasible and reliable tools combining measurements of growth, health, physical and cognitive function. Previous studies have individually shown that poor linear growth is associated with reduced cognitive development^17^, grip strength^18^ and increased cardio-metabolic risk factors^19^. However, the absence of holistic assessments combining neurodevelopment, physical fitness and growth has led to a call for further studies measuring a range of outcomes^13 20^. Previous epidemiological work has identified height at 2 years as a strong predictor of later human capital^21^. However, assessing the efficacy of early-life interventions requires long-term follow-up to measure later functional outcomes. A comprehensive assessment is vital to understand school-age trajectories across different functional domains and the impact of risk and protective factors and potential interventions.

To overcome these gaps, we developed an integrated test battery combining measures of school-age growth, body composition, physical and cognitive function in sub-Saharan Africa. Our toolbox has been specifically designed for use within low-resource communities using portable equipment to make testing highly accessible. Here, we use this new test battery to evaluate the associations between growth, physical function and cognitive functional domains in a cohort of Zimbabwean children.

## METHODS

A conceptual framework was developed which identified hypothesised relationships between a child’s environment, growth, physical and cognitive function (Figure 1a). The School-Age Health, Activity, Resilience, Anthropometry and Neurocognitive (SAHARAN) toolbox (Figure 1b) was designed to test these relationships, using a stepwise approach described in detail in Supplementary Information^22^.

**Fig 1a:**
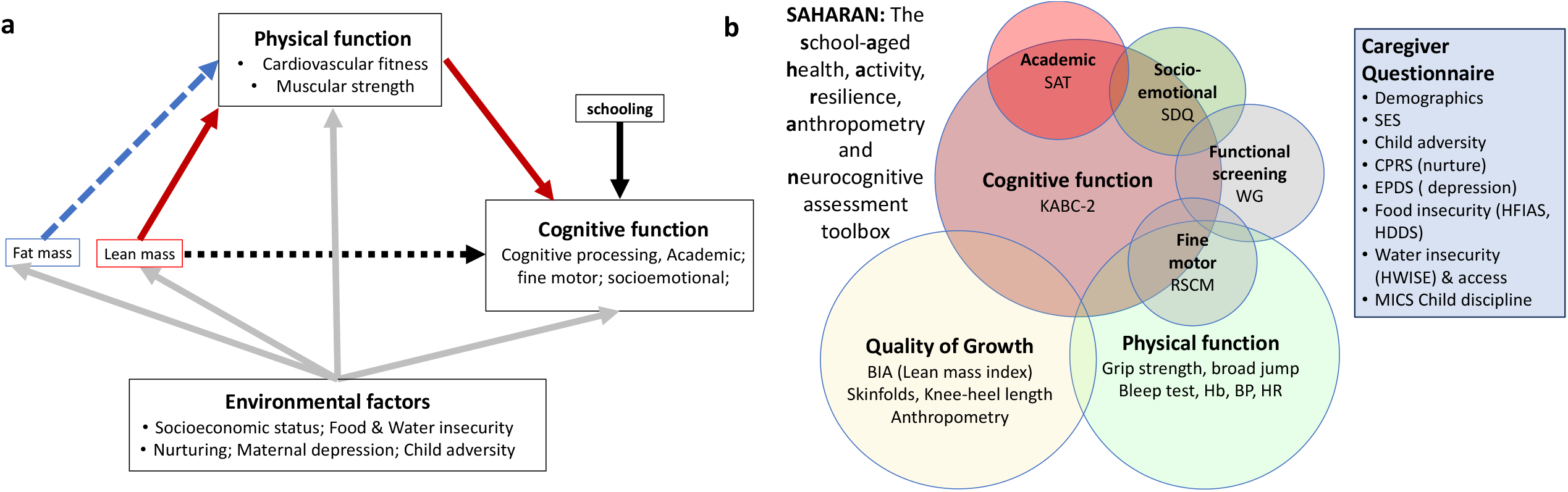
Conceptual framework to inform the choice and design of tools. Fig 1b: The SAHARAN toolbox: A 4-hour assessment comprising child growth, body composition, cognitive and physical function assessment with simultaneous caregiver questionnaire. KABC-2: Kaufmann Assessment Battery for Children 2^nd^ edition, SDQ: Strength and difficulties Questionnaire, WG: Washington Group / UNICEF Child Function module, RCSM: Rapid continuous sequential movements, BIA: Bioimpedance analysis, Hb: Haemoglobin, BP: Blood pressure, HR Heart rate, SES: Socioeconomic status, CPRS: Child-parent relationship scale, EPDS: Edinburgh postnatal depression score, HFIAS: Household Food Insecurity Assessment Scale, HDDS: Household Dietary Diversity scale, HWISE: Household Water Insecurity Experiences Scale, MICS: Multi-indicator cluster survey (UNICEF) for Discipline Questionnaire

In brief, we first conducted a detailed literature review, which identified recent systematic reviews describing tools for measuring cognitive function^13 23 24^, a recent toolkit published by the World Bank^25^, and a test battery for physical function^26 27^. Second, individual assessment tools were screened for their use in LMICs^28 29^ using selection criteria based on the COSMIN tool (see Supplemental Material)^30^. Third, a range of international child development, nutrition and sports science experts were contacted to provide input on each tool, including their content validity and applicability for a rural sub-Saharan African context^31^. Fourth, pre-testing, cognitive interviewing and piloting of instruments led to further adaptation and selection of tests for the final toolbox (Figure 1B). Fifth, detailed standardisation exercises were conducted to assess intra- and inter-rater reliability. Finally, construct validity and internal consistency were assessed by examining the relationships between different tests within similar cognitive domains (convergent validity), as well as relationships between domains.

### Tests selected for the SAHARAN toolbox

Detailed descriptions of the final test battery are provided in Supplementary materials. Briefly, the SAHARAN toolbox comprises a single assessment conducted with the child and caregiver, focused on three domains: growth and body composition, physical function and cognitive function (Fig 1b). Growth is assessed using standard anthropometry (weight, height, head, arm, waist, hip and calf circumferences). Body composition is assessed using bioimpedance analysis (BIA) to estimate lean mass^32^, skinfold thicknesses to measure subcutaneous fat, and knee-heel length to examine relative body proportions^33^. For physical function, handgrip^18^ and the broad jump^18^ measure strength, and the shuttle run test (SRT) assesses cardiorespiratory function^34^, with additional measurements of blood pressure, heart rate and haemoglobin. Initial blood pressure values measured electronically were systematically high, so manual blood pressure readings were introduced for the last 18 children. Cognitive function is measured using the mental processing index (MPI), calculated from 8 core subtests of the Kaufmann Assessment Battery for Children (KABC-2)^29 35^. Academic function is assessed by the school achievement test (SAT) which measures reading, writing and numeracy. During piloting it was adapted by selecting the appropriate font and choice of letters for reading that were common in Shona and Ndebele, as well as the addition of reading syllables. Fine motor function is evaluated by the time to complete sequential finger tapping^36^, with a shorter time representing improved fine motor function. Socioemotional function is measured by the Strengths and Difficulties Questionnaire^37^ and a short socioemotional questionnaire asked directly to the child. Child disability and overall caregiver-reported function are measured using the Washington Group / UNICEF Child Functioning Module^38^. A detailed caregiver questionnaire measures household demographics, adversities^39 40^, socioeconomic status^41^, food security^42 43^, water security^44^, and caregiver nurturing^45^, depression^46^ and discipline ^47 48^.

### Participant and public involvement

School-age participants and their caregivers were not involved in setting the research question or outcomes. However, they were involved in design and implementation of the questionnaires and tools. All tests to measure school-age child growth, physical and cognitive function were discussed with the District Health Executive, community leaders, and health centre committees in Zvambande and Makusha. All questionnaires underwent cognitive interviewing with community members who suggested alterations and feedback. Tools were then pre-tested with local families to ensure acceptability, with feedback sought from children and caregivers. Extensive sensitisation of the communities was undertaken by the study team and village health workers in conjunction with screenings of a community-made film which explored their previous experiences of being involved in research. During sensitisation and consent visits, it was explained that findings from the current study would not be shared because the aim was to assess the feasibility and acceptability of the SAHARAN toolbox.

### Study site and data collection

This study was conducted in Zvamabande (rural) and Makusha (peri-urban) regions of Shurugwi district in Midlands Province, Zimbabwe. During sensitisation events, the proposed tests were explained to communities. Eligible children aged 7 years were identified by village health workers (VHW), then 80 children were randomly selected by computer, and a sensitisation visit to the family was conducted by the VHW. If the family expressed interest in participating, a date was agreed for a visit.

Assessments were undertaken in the community using portable equipment (Supplementary Fig S1). A handwashing station was erected and facemasks distributed, in line with district COVID-19 policies. One or two tents were pitched close to the household with four folding chairs, where the mother and child could see each other at all times. The child cognition measurements were administered in the tent using a folding table. Data were collected using Open Data Kit (ODK)^49^ on tablet computers (Samsung Galaxy Tab A) for most measurements, enabling appropriate data skips and plausibility checks; some cognition measurements used paper forms (KABC-2, School Achievement Test).

### Data analysis

Growth measurements were converted to Z-scores using WHO reference standards^50^. BIA data were converted into lean mass index, defined as 1/Z (the average impedance), which is independent of height^51^. The ‘impedance index’ defined as height-squared divided by average impedance (H^2^/Z), was also calculated as a direct estimate of relative lean mass; this incorporates height in the estimation, as lean mass always scales strongly with height^52^. Impedance index therefore acts as a composite marker of muscle and organ mass relative to height. Phase angle was used as a marker both of cell mass and tissue health^53 54^. Least squares linear regression was used to explore associations between components of the SAHARAN toolbox using Stata Version 15 (StataCorp LLC, College Station, TX). This explored the relationships between domains of growth and function.

### Ethics

Caregivers gave written informed consent and children gave written assent to participate. Ethical approval was provided by the Medical Research Council of Zimbabwe.

## RESULTS

Of 180 children identified by village health workers, 23 were excluded because they were outside the study age range, leaving 157 eligible children, from whom 80 were randomly selected. Among these 80 children, 3 children had the wrong age on documentation (birth certificate or health card) and were excluded, and 2 families were not interested in joining; 5 random replacements were therefore identified. Overall, 80 children (39 girls; 49%) were enrolled and underwent assessments between September 3^rd^ and December 4^th^ 2020.

Baseline characteristics are shown in Table 1. Children had a mean age of 7.6 years (SD 0.2). Most households (79/80; 99%) undertook subsistence farming. Children had a mean of 3.1 years (SD 0.7) of prior schooling, with girls having more previous school exposure than boys. The impact of COVID-19 was evident: only 15/80 (19%) children had enrolled in school during that academic year, and all caregivers reported their child missing school due to COVID-19 restrictions. All children completed the full battery of tests and questionnaires during a single visit, which lasted 4-5 hours.

**Table 1:** Baseline characteristics of participants, household and selected adversity factors in the sample. HAZ: Height-for-age Z-score, WAZ: Weight-for-age Z-score, EPDS: Edinburgh postnatal depression score^46^, HFIAS: household food insecurity assessment scale^55^, HDDS: Household Dietary diversity scale^43^, HWISE: household water insecurity experience scale) ^44^.

| **Child characteristics** | **All (N=80)** |
| --- | --- |
| Female, N (%) | 39 |
| Age, yrs; mean (SD) | 7.6 (0.2) |
| HAZ; mean (SD) | -0.63 (0.8) |
| Stunted (HAZ<-2), N (%) | 2 (3%) |
| WAZ; mean (SD) | -0.55 (0.8) |
| Underweight (WAZ <-2), N (%) | 3 (4%) |
| BMI-Z score, mean (SD) | -0.27 (0.9) |
| Enrolled in school, N (%) | 15 (19%) |
| Reported schooling at home, N (%) | 33 (41%) |
| Years in school; mean (SD) | 3.1 (0.7) |
| **Caregiver characteristics** |  |
| Mother; N (%) | 54 (68%) |
| Grandmother; N (%) | 20 (25%) |
| Other; N (%) | 6 (7%) |
| Age, years; mean (SD) | 39.8 (11.7) |
| Years of schooling; mean (SD) | 9.1 (2.7) |
| Married, N (%) | 63 (79%) |
| EPDS score; mean (SD) | 6 (5) |
| **Household characteristics** | **n** |
| Household size; median (IQR) | 3 (2,5) |
| Electricity to home; N (%) | 9 (11%) |
| Drinking Water > 5 mins walk; N (%) | 62 (78%) |
| Own radio; N (%) | 59 (74%) |
| Own phone; N (%) | 79 (99%) |
| Own TV; N (%) | 36 (45%) |
| Own car/truck; N (%) | 8 (10%) |
| Own solar panel; N (%) | 62 (78%) |
| Own 2 or more children's books; N (%) | 29 (36%) |
| HFIAS median score (IQR) | 6 (1, 10) |
| HDDS median score (IQR) | 8.5 (7,10) |
| HWISE median score (IQR) | 0 (0,1) |
| **Adversity characteristics** |  |
| Death in household | 26 (33%) |
| Adult sick / injured > 3 mo | 11 (14%) |
| 1 crop failure | 60 (75%) |
| 2 or more crop failures | 37 (46%) |
| Loss of family possessions | 27 (34%) |
| Alcohol problem | 16 (20%) |
| Debt problem | 16 (20%) |
| >2 Child hospital admissions | 2 (3%) |
| Business failure | 40 (50%) |
| Lost job | 22 (28%) |
| Lost job > 6 mo | 15 (19%) |
| Child separation > 3 mo | 17 (21%) |

### Growth and body composition

Body composition and anthropometry measurements were well tolerated by all children. Mean (SD) height-for-age and weight-for-age Z-scores were −0.63 (SD 0.81) and −0.55 (0.85), respectively, and were similar by sex; 2/80 (3%) children were stunted and 3/80 (4%) were underweight. The mean body mass index (BMI) was 15.3 kg/m^2^ (SD 1.4, range 12.5 to 21.6). WAZ was significantly associated with increases in head circumference, HAZ, lean mass index (Fig 2a), phase angle, knee-heel length, impedance index and skinfold thickness (Fig 2b, supplementary table S6). By contrast, HAZ was only associated with impedance index and knee-heel length and not with lean mass index (Fig 2c) or skinfold thickness (Fig 2d, table S6). Increases in phase angle were associated with increases in impedance index and lean mass index (table S6). There was no association between total skinfold thickness (measuring subcutaneous fat) and bioimpedance measures of lean mass including impedance index, lean mass index (Fig 2e) or phase angle (Table S6).

**Figure 2:**
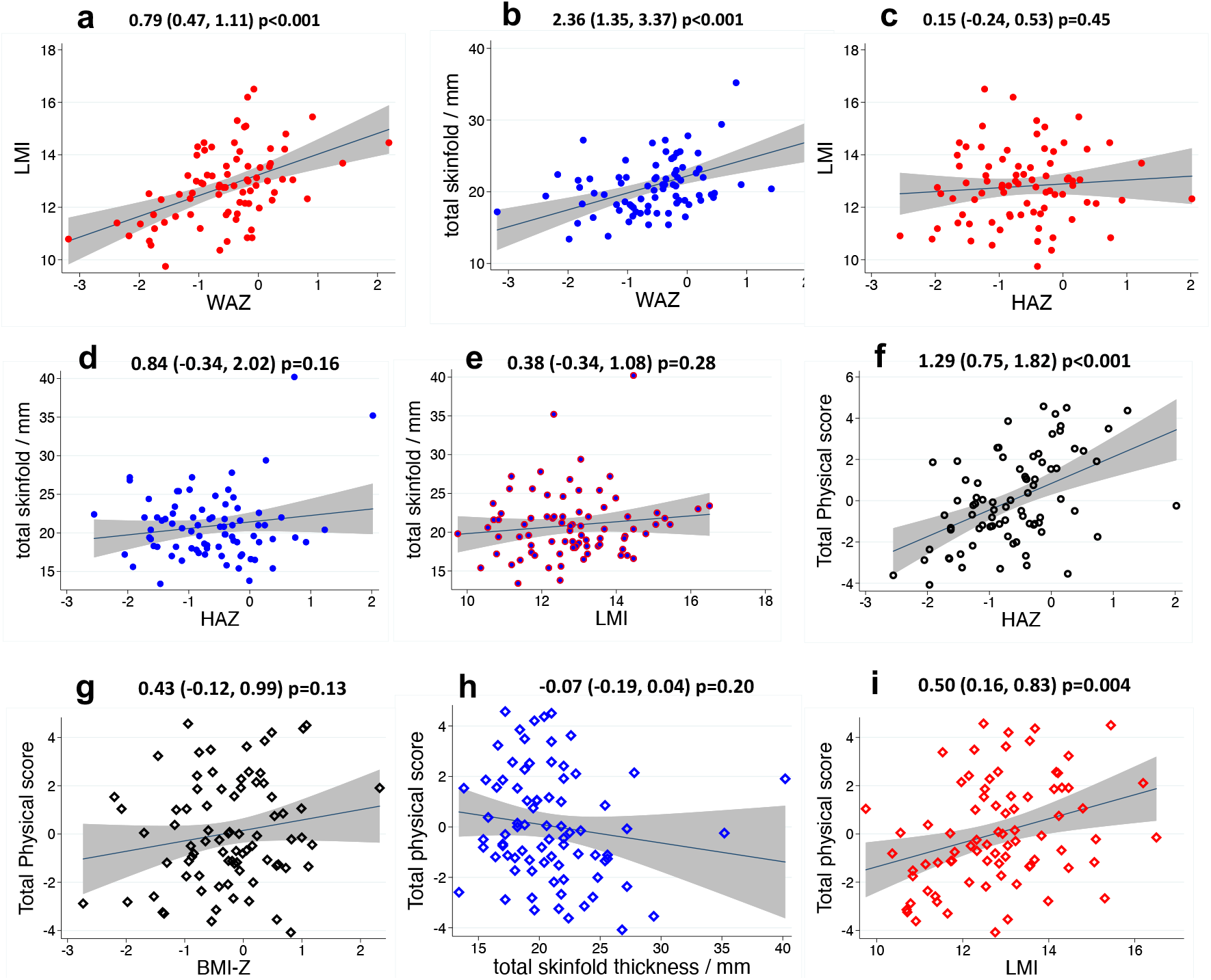
SAHARAN toolbox results describing growth, body composition and their association with physical function. a: Lean mass index (LMI) was highly associated with weight-for-age Z-score (WAZ), due to increasing lean mass with weight. b: Total skinfold thickness was strongly associated with WAZ since fat mass increases with weight. c: LMI was not associated with height-for-age Z-score (HAZ) as LMI adjusts for the contribution of height to lean mass. d & e: Total skinfold thickness was not associated with HAZ or LMI. f: Total physical function score (TPS) was highly associated with increasing HAZ. g: TPS was not associated with Body Mass Index Z-score (BMI-Z) because of differing contributions from both fat and lean mass. h: TPS was not associated with skinfold thickness, with a possible trend suggesting skinfold thickness may negatively contribute to total physical function. i: TPS was strongly associated with LMI showing the positive contribution of lean mass to physical function independent of height.

### Physical function

Physical function measurements were well tolerated. All tests were highly associated with growth measurements (Supplementary table S7, Fig 2). For every unit rise in HAZ (∼6cm^50^) or WAZ (∼3.5Kg^50^), maximum grip strength, broad jump, and shuttle run test (SRT) level all increased significantly. Total physical score was calculated by adding the standardised scores from the 3 physical function tests (handgrip strength, broad jump distance and level reached in the shuttle run test).

The total physical score (TPS; defined by adding standardised scores for the 3 physical tests), was significantly associated with HAZ (Fig 2f), WAZ, hip circumference, MUAC and calf circumference (Table 2). TPS was only weakly associated with Body Mass Index Z-score (BMI-Z) because of differing contributions from both fat and lean mass (Fig 2g). Measures of fat mass such as skinfold thickness (Fig 2h) and waist circumference (Table S7), were not associated with physical function. By contrast, lean mass index (Fig 2i), impedance index and phase angle were highly associated with physical function (Table S7). The differential contributions of measures of body composition are summarised in Figure 4. Children had a mean haemoglobin of 126 g/L (SD 10.1); 5 children (6%) had measurements below the WHO anaemia threshold of 110 g/L (lowest value 96 g/L), but no child had symptomatic anaemia. Haemoglobin was not associated with any functional score.

**Figure 3:**
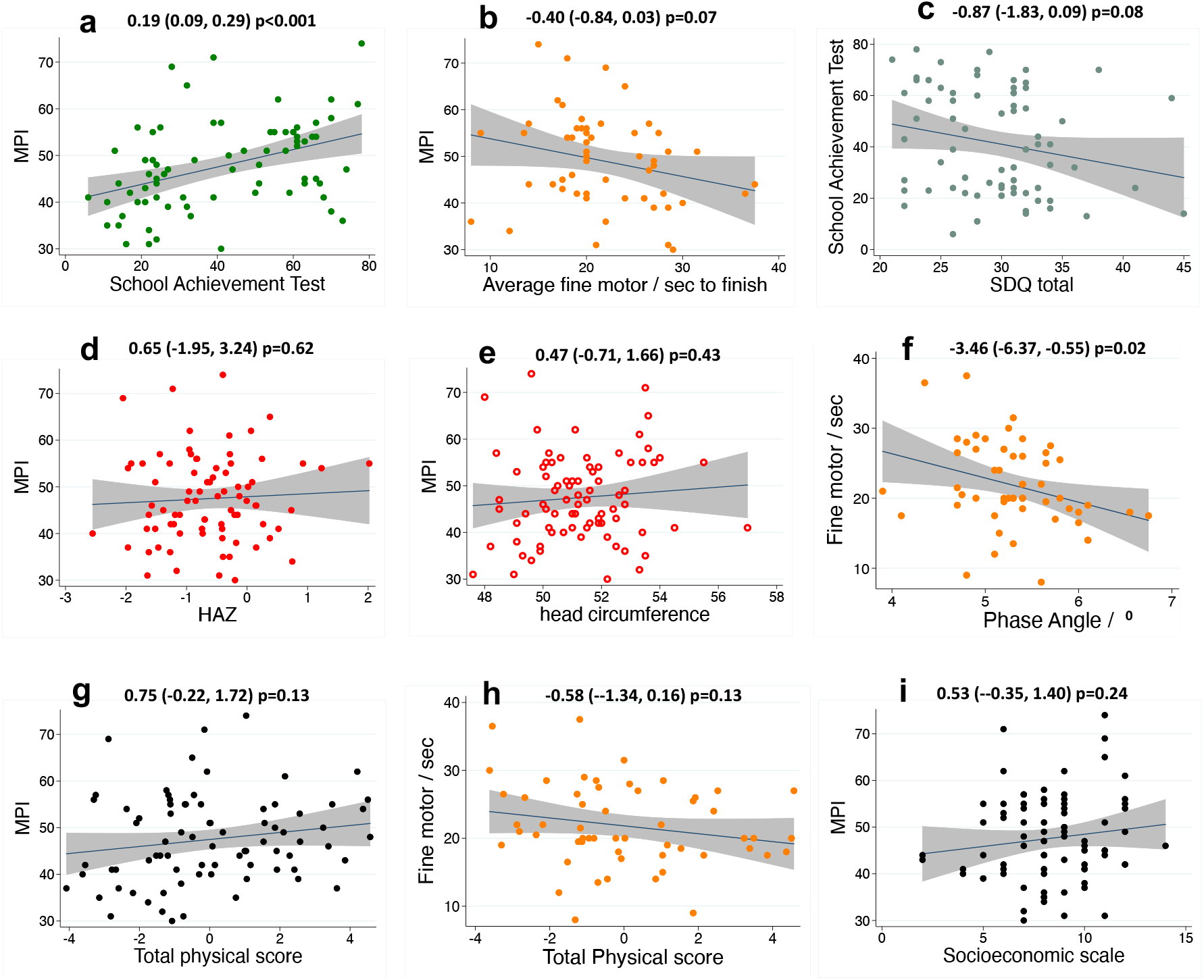
SAHARAN toolbox results describing cognition results and their association with growth and physical function. a: Internal consistency showed MPI (Mental processing index) was associated with increasing School achievement test (SAT). b: Higher MPI was associated with faster fine motor completion time. d: Lower school achievement test scores were weakly associated with reduced socioemotional function, as measured by a higher strengths and difficulties score (SDQ). d & e: MPI was not associated with growth parameters of HAZ or head circumference. f: Faster fine motor function was associated with increased bio-impedance phase angle (a marker of cellular health and membrane quality). g & h: TPF was not associated with MPI or fine motor function, although plausible potential trends were observed. i: MPI was not significantly associated with socioeconomic scale.

**Figure 4:**
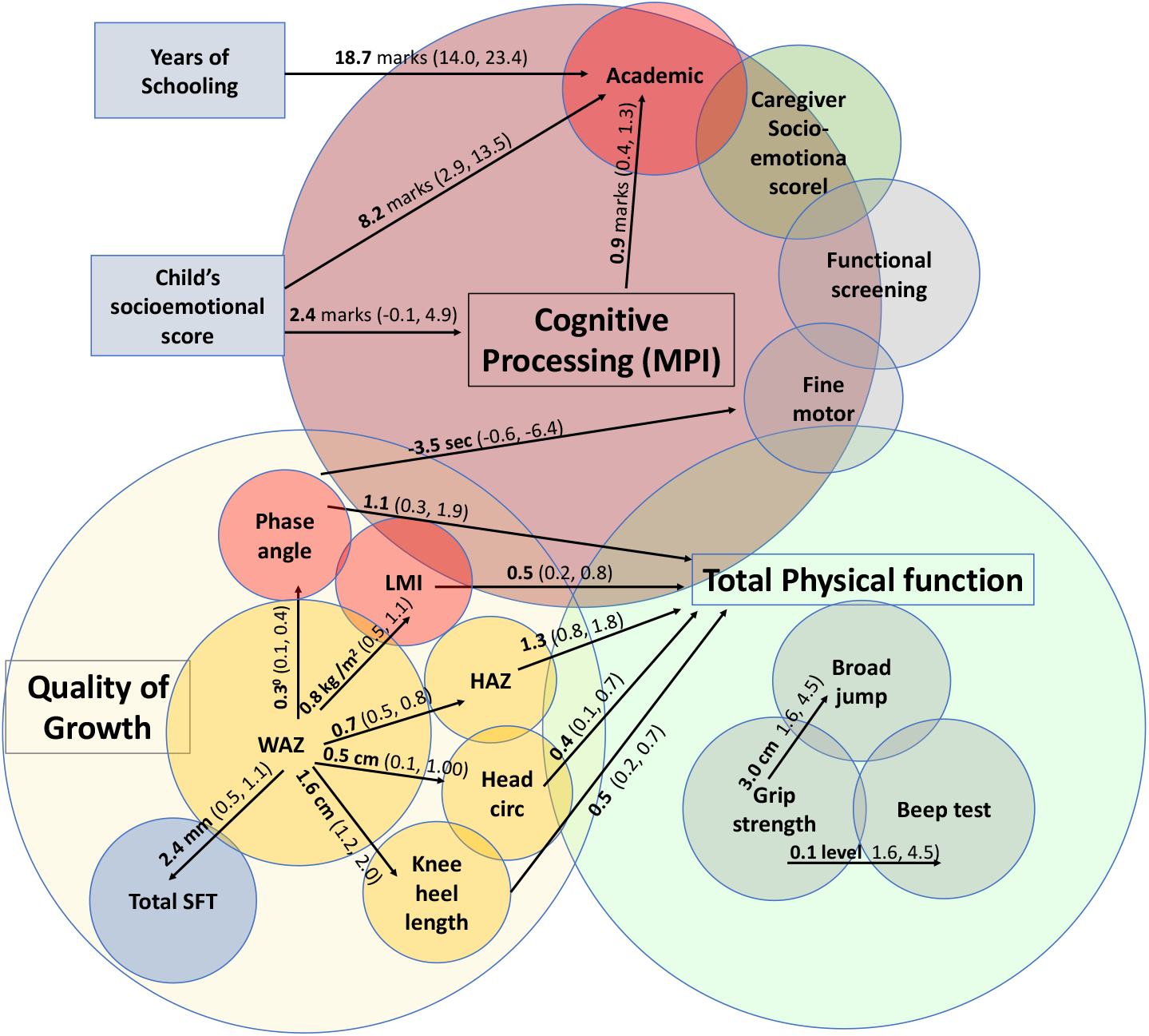
Significant associations within the SAHARAN toolbox between quality of growth, total physical function and cognitive function domains. Linear regression coefficients and confidence intervals are shown.

### Cognition results

The School Achievement test (SAT) was performed in 73 (91%) children (see Supplementary Materials); in the first 7 children a Shona version of the Early Grade Reading Assessment (EGRA) was performed, but scores on this tool were extremely low due to a floor effect: the tool was too advanced for the participants tested. Generally, the SAT provided a good range of variability; 18 (23%) children were unable to recognise any letters and 9 (11%) were unable to write any letters.

Direct measurements of cognition showed internal consistency. The Mental Processing Index (MPI), which provides a total cognition score from the KABC-2, was strongly associated with the SAT (Fig 3a) and weakly associated with fine motor skills (Fig 3b). However, socioemotional function as measured by the caregiver-reported SDQ was only weakly associated with SAT (Fig 3c) and not with MPI or fine motor function (Table S8). There was strong evidence that each additional year of schooling was associated with the SAT, and weak evidence for a relationship with the MPI; there was no association with fine motor speed or SDQ score (Table S8). The child’s socioemotional questionnaire was strongly associated with the SAT, and there was weak evidence for a relationship with the MPI score; there was no relationship between the child-reported socioemotional score and the caregiver-reported SDQ (Table S8).

There were no significant associations between cognitive test scores and measures of growth such as HAZ (Fig 3d) or head circumference (Fig 3e). A higher phase angle was significantly associated with a shorter fine motor completion time (representing faster fine motor function) (Fig 3f). The total physical function score was not associated with MPI (Fig 3g) or quicker fine motor function (Fig 3h). These associations are also summarised in figure 4.

### Caregiver questionnaire

Socioeconomic scale was positively associated with the Household Dietary Diversity Score (HDDS). For every unit rise in HDDS, the socioeconomic score increased by 0.46 (95%CI 0.23, 0.69, p<0.001). For the household food insecurity assessment scale (HFIAS), every unit rise in HFIAS (which corresponded to worse food insecurity) was associated with a decrease in the socioeconomic score of −0.11 (95% CI −0.19, −0.03, p=0.01). Socioeconomic score was not associated with cognitive processing as measured by the MPI (Fig 3i). Generally, there was little documented water insecurity so HWISE scores were low (Table 1). 10/80 (13%) caregivers screened positive for clinically significant depression, but this was not related to socioeconomic status or any test scores in the child. Measures of nurturing such as the Child Parent Relationship Scale (CPRS) and child discipline score were not associated with cognitive, growth or physical function in the child.

No child had serious disability. Three children (4%) had parent-reported problems in learning, but this was not associated with cognitive performance. The adversity questionnaire documented a high prevalence of adversities over the 7 years since the child’s birth, with a mean of 5 (SD 3.1) events; only one household reported no adversities and three reported only one adversity. The most common adversities were crop failure (N=60; 75%), business failure (N=40; 50%), lost family possessions due to hardship (N=27; 34%), household death (N=26; 33%), job loss (N=22; 28%), at least one household member with an alcohol problem (N=16; 20%), worries due to debt (N=16; 20%) and more than 6 months of unemployment (N=15; 19%). Of the 80 children, 17 (21%) had been admitted to hospital; only 3 children (4%) had been admitted to hospital twice or more. The total adversity score had no association with growth, physical or cognitive function.

Taken together (Figure 4), the SAHARAN toolbox showed significant associations between growth measures, which were also strongly associated with physical function. Cognitive function measurements showed internal consistency and significant associations with schooling exposure and child socioemotional score. Growth and physical function were not associated with cognitive function, except for phase angle which was associated with fine motor function.

## DISCUSSION

There is increasing recognition of the need to assess child health, growth and function together at school-age, to more holistically evaluate long-term outcomes following early-life adversities and interventions. We formulated a conceptual framework then developed and deployed the SAHARAN toolbox to investigate the relationships between these domains in a cohort of school-age Zimbabwean children. We found clear associations between linear growth and physical function across a range of tests evaluating muscle strength and cardiovascular fitness, highlighting the effect of attained height on physical performance. The quality of growth in early life was also associated with school-age outcomes, since lean mass index was related to physical function, independent of height. Finally, we found associations between different domains of directly measured school-age cognitive function, including global cognition, academic and fine motor skills. This highlights the interconnected aspects of school-age child development, with the potential for interventions to influence multiple cognitive domains. Collectively, our data demonstrate the feasibility and utility of combining growth, body composition, physical and cognitive function to more comprehensively unravel how early-life conditions shape prioritization and trade-offs in later function. Our novel, holistic assessment battery can now be applied to larger cohorts to gain a richer understanding of school-age health outcomes in LMIC settings.

We found significant and intriguing associations within and between growth and physical function measures (Figure 4). Child height was strongly associated with all the physical tests. Height has previously been strongly correlated with grip strength at school-age^56^ as well as with strength of other muscles^57^. The strong correlations between different muscle function tests indicates a global effect of stature (possibly also mediated by bone growth) on whole-body muscle strength^58^. Height is closely associated with weight, but the association of weight with the shuttle-run test was weaker than for height, since the SRT measures cardiovascular fitness. Neither BMI nor waist circumference were associated with physical function, despite these measurements generally being recommended for body composition in adolescents and adults^26^. This is probably because of the narrow BMI range at 7 years of age. Therefore, more detailed body composition measurements provided further insight into the proportion of fat and lean mass, which were closely correlated with function. MUAC, hip circumference and calf circumference were all associated with physical function, as they all measure muscle as well as subcutaneous fat. When lean and fat mass were measured individually by more advanced body composition measurements, increasing lean mass index was highly associated with total physical function. By contrast, fat mass represented by skinfold thickness was not significantly associated, and a possible trend suggested the opposite relationship. This is expected as lean mass increases with both strength and fitness^59^. Similarly, fat mass has previously been shown to have no relationship to grip strength^56^. Indeed, muscular strength is inversely associated with increasing adiposity in children and adolescents^60^. Taken together, these data provide intriguing evidence that promoting early-life linear growth and lean mass accretion may help to improve later physical function and reduce the risk of chronic disease^60 61^, supporting trends observed globally^9^.

Cognitive tests demonstrated expected associations with each other and with schooling exposure, which had been negatively impacted by the COVID-19 pandemic. The SAT score was significantly associated with years of schooling, while the mental processing index, derived from the KABC-2 tests, had a less strong association with prior schooling; this is perhaps unsurprising, since the KABC-2 is designed to include novel subtests that children would not be exposed to in schools^35^. The trend towards improved fine motor skills with higher MPI is also consistent with the correlation between different domains of cognition. The fine motor tool we used has not previously been directly compared with cognition measures, although stunted compared to non-stunted Jamaican children were reported to be approximately 3 seconds slower on sequential finger tapping ^36^.

Each additional point on the child socioemotional score (representing the child reporting feeling happier and more supported at home) was associated with additional marks on the SAT and the MPI (Fig 4). This suggests that the child’s perception of their home environment can influence their academic performance, providing some early evidence of novel areas for psychological support. By comparison, there was a weaker association between the Strengths and Difficulties Questionnaire (SDQ) and the SAT. This may partly be because SDQ is a caregiver-reported tool, although caregiver-reported and teacher-reported SDQ were associated with academic scores among children in Brazil^62^. A recent follow-up of the MAL-ED cohort showed that socioeconomic status was associated with cognitive scores at 5 years^63^; our study found a possible trend towards increasing MPI with a rise in socioeconomic scale.

When comparing growth parameters and cognitive function, the only significant association was between increased phase angle and improved fine motor function (Fig 3f). This is plausible since higher values of phase angle are thought to reflect improved cellular health, particularly membrane function^64^. None of the other growth or body composition measurements were associated with any cognitive test. This was largely expected, as a recent systematic review highlighted that growth is not a reliable proxy for cognitive function^13^, which emphasises the importance of direct measures of cognition at school age. There is some evidence to suggest acute exercise before cognitive testing can improve cognitive performance^65^, hence we measured cognitive function routinely before physical function. Two systematic reviews have suggested a positive effect of physical activity on cognitive function but with variable and inconsistent evidence^66 67^. Subsequent studies have shown moderate correlation in direct measurement^68^, whilst two randomised trials of an exercise intervention have shown no effect on cognitive performance^69 70^.

Our study has several strengths. Firstly, the SAHARAN toolbox is the first community-based, school-age assessment battery to combine measurements of school-age growth, body composition, physical and cognitive function together with a questionnaire on contemporaneous environmental factors. We included several novel tools, including sequential fine motor tapping and the use of electronic data collection in a rural setting. The toolbox was portable, and successfully deployed during the COVID-19 pandemic, providing reassurance that it can be used in other settings. Our study also had several limitations. First, we enrolled a convenience sample of children for this study, and our findings may not be applicable to the most hard-to-reach children in all communities. However, we enrolled children from both peri-urban and rural areas, where poverty and food insecurity are both pervasive, meaning our results may be applicable to similar settings. The 4-hour test battery limits the number of children who can be measured and is a substantial time burden for families. However, future factor analyses will be undertaken to shorten the SAHARAN toolbox into the most discriminatory cognitive and non-cognitive metrics for greater applicability. The KABC-2 does require considerable training, though in this study it was conducted online by experienced trainers. With permission, we adapted two KABC-2 subtests which used Western images and concepts unfamiliar to rural African children. Finally, this cross-sectional study only measured a single time-point, so any associations with improvements or declines in function over time could not be detected. For example, a higher adversity score may have a much greater impact in early life, which may not be detected in this cross-sectional study.

In summary, we designed a holistic toolbox to fill a much-needed gap during the ‘missing middle’ of childhood when child health outcomes are generally overlooked. This assessment battery provides a novel combination of growth, body composition, cognitive and physical function measurements for school-age children, which would be applicable in research trials and programmes globally. Results from this initial study show consistency between the different cognitive measures used, and intriguing associations between growth, body composition and physical function. Our findings highlight the importance of early-life growth and relative lean mass for improving physical function and reducing chronic disease risk^61^. This study reaffirms the value of combining assessments of body composition, physical and cognition function, and provides the opportunity to now characterise the effects of early-life exposures and interventions on school-age growth and development in multiple settings.

## Data Availability

Supplementary material will be added.

## ACKNOWLEDGEMENTS

We thank Zvitambo staff for their help with this study, particularly Virginia Sauramba and Sandisiwe Ndlovu (compliance), Stephen Moyo and Peter Mapuranga (logistics), Theo Chidawanyika (IT) and Bernard Chasekwa for statistical assistance with the socioeconomic scale. We thank Prof Jean Humphrey for valuable discussions in planning this study. We thank Jackie Namukooli and Mary Nyakato for initial KABC-2 training and Prof Tamsen Rochat, Samu Dube and her team for further online training for the KABC-2. We thank Prof Susan Chang-Lopez for assistance with sequential finger tapping. We also thank Dr Marko Kerac and Dr Keith Brazendale for helpful discussions regarding the physical function tests, and Dr Natasha Lelijveld and Dr Carlos Eternod-Grijalva for general advice on the SAHARAN design.

