## Supporting Methods for "Characterising school-age health and function in rural Zimbabwe using the SAHARAN toolbox"

### Supplementary Material

#### Outcomes and constructs of interest

The SAHARAN toolbox combines measures of growth, body composition, cognitive and physical function to provide an overall assessment of child function. This new combined tool was designed to enable us to identify relationships between domains which are not usually measured simultaneously. Here we describe the different constructs of interest and the choice of measurement instruments used, based on the COSMIN study design checklist, and the methodology used for pre-testing and piloting, following principles previously described[1].

##### 1. School-aged Cognitive function

Cognitive function is best assessed using multiple tools that measure distinct domains. Cognition is affected by multiple factors including undernutrition, nurturing, maternal and child interactions and stimulation[2]. Children in low-resource settings often have multiple exposures including socioeconomic adversity, undernutrition and lack of psychosocial support[3]. These may have differential impacts on cognitive domains, reinforcing the need to measure multiple domains, as described below.

###### 1.1 Child functional summary

A measure of overall child function can determine difficulties in sight, hearing and mobility, as well as socioemotional areas including behaviour, learning and psychological functioning[4]. By focusing on what the child may have difficulty doing, this provides an overview that is complementary to specific areas of cognitive function.

##### 1.2 Cognitive processing

Traditional intelligence testing methods relied on measuring *acquired knowledge*, such as intelligence quotient (IQ) testing. These tools were frequently developed in Western settings with high school enrolment rates: their use in low-resource settings is contentious, particularly in areas with variable schooling exposure [5]. Therefore, tools that measure *learning potential* are often preferred in low-resource environments[5]. These measure 'cognitive processing', which encapsulates the underlying processes and skills necessary to solve tasks, and are less sensitive to cultural and schooling bias[5 6]. Hence a tool that measures cognitive processing across several domains of short and long-term learning memory, spatial reasoning and problem solving is preferred.

##### 1.3 Academic skills

Complementary to cognitive processing, traditional measures of educational achievement are also important predictors for later academic and socioemotional function[7]:

i) Literacy: Early measures of language are indicative of future function including literacy, behaviour and socioemotional function[8]. Unless children learn to read at an early age, they cannot absorb more advanced skills and content that rely on reading, and typically fall behind in educational achievement[9]. Similarly, writing skills such as the child's name[10] or dictation of words have also been demonstrated to be discriminatory for early-grade schooling exposure[11] and emergent literacy skills[10].

ii) Mathematical skills: Mathematical literacy (including numeracy) is increasingly recognised as a core skill in everyday life[12]. Hence measuring early-age mathematical capabilities similarly predicts later engagement, learning and educational achievement[13].

##### 1.4 Executive function

Executive function includes universal skills such as inhibitory control, working memory, attention, and cognitive flexibility[14]. These skills enable children to plan, focus attention, set and achieve goals. They are highly related both to school readiness in young children[15] and later educational outcomes[16]. Executive function is also related both to socioemotional function[15] and academic skills[7 15]. These goal-directed behaviours may be viewed as culturally universal skills[17] and therefore a tool that can specifically measure executive function may be included in future measurements.

##### 1.5 Behaviour and Socioemotional function

Behavioural and emotional problems are common in children. Externalising behavioural problems such as attention deficit hyperactivity disorder may occur in up to 9% of preschool children in high-income settings[18] and this may be higher in some low-resource settings[19]. Child mental health (including behavioural measures) is increasingly recognised as an important measure affecting cognitive function[20]. Cognitive, academic, executive and socioemotional function are inter-related both in early childhood[21] and adolescence[22]. Therefore, this domain is also included and often measured through interview of caregivers or teachers[19 23].

##### 1.6 Fine motor function

Fine motor coordination is important for completion of many tasks, and has previously been identified to be reduced in stunted children, particularly those with worse academic function[24]. Reduced ability in fine motor function has also been associated with low birthweight and reduced socioemotional function[25], as well as reduced academic function[26]. This may be in part due to lags in development[27].

.

#### 2. School-aged physical function

Systematic reviews have shown that child physical function and health are best assessed by combining separate measures of cardiovascular fitness, muscular strength and body composition[28 29]. For example, cross-sectional data have shown that both cardiorespiratory fitness and muscular strength in children are associated with lower cardiovascular disease risk factors in later life (including lower blood pressure, blood lipids and adiposity)[28 30]. There is also strong evidence that changes in muscular strength from childhood to adolescence are negatively associated with changes in overall adiposity[28]. Therefore, physical function measurements should include both cardiovascular fitness and muscular strength.

The physical function tests selected were similar to the ALPHA[29] and PREFIT[31] test batteries. The choice of tests was based on a systematic review of the literature,[32] combined with what had been previously piloted in LMIC (Kerac, personal communication). There is increasing evidence of physical activity being associated with improved cognitive ability, but this is mainly in higher income settings[33]. Physical fitness has also been described as a powerful marker of child health and future cardiovascular risk[30]. With the double burden of malnutrition, there is a rising trend of obesity and overweight children in sub- Saharan Africa, and increasing interest in methods to measure physical fitness[34].

#### 3. Growth

There is emerging evidence that stunted growth in early life affects later height, body composition and physical parameters such as blood pressure[35 36]. Body composition at school-age and adolescence (measured using body mass index, skinfold thickness and waist circumference) is associated with cardiovascular risk factors such as blood lipids and carotid artery narrowing[28]. There is also strong evidence indicating that a high BMI in childhood and adolescence increases the risk of death later in life[28]. A recent review highlighted that interventions that solely improve linear growth, are only weakly associated with cognitive function[37]. There is some intriguing emerging evidence that body composition at birth is associated with socioemotional function, with increasing lean mass improving socioemotional health and fat mass decreasing it[38]. This likely reflects maternal life history trade-offs among offspring[39]. Therefore growth measurements included both anthropometry and body composition.

Table S1: Tool selection

| Domain | Sub-domain | Tool | Outcome(s) | Validity | Reliability | Responsiveness |
|---|---|---|---|---|---|---|
| Cognitive Function | Cognitive Processing | Kaufmann Assessment Battery for Children (KABC2) | KABC2 measures across four domains of sequential, planning, learning and simultaneous scales.<br>8 core subtests can be combined as the mental processing index (MPI) to provide a global outcome | The KABC2 was originally developed and validated using a large sample in the USA[40]. It has since been widely used across Africa[41], demonstrating robust factor analysis in Uganda[42] and psychometric validity in rural South Africa[43]). The KABC2 group of cognitive tests show less bias to school exposure in low-income settings [43]. | Reliability has been demonstrated in USA[40] and South Africa[44]. Recently, the QualiND model has demonstrated improved KABC-2 monitoring and quality assurance using regular video review[45] across multiple countries and languages in Africa, including with a Shona translation in Zimbabwe[45] | Using KABC-2, a significant effect on cognition was detected with a nutrition intervention in South African children aged 6-11 years on two of the subtests[46] whilst in Ethiopia, 5 year olds with poorer growth also had worse KABC-2 scores[47]. HIV positive children performed significantly worse than HIV-negative children in South Africa, Zimbabwe, Malawi and Uganda [48]. Similarly in separate studies in Burkina Faso, both stunted children [49] and those exposed to alcohol in pregnancy[49] performed significantly worse on KABC-2 subtests. |
| Cognitive Function | Executive Function | Plus EF[1] | Inhibitory control: Hearts and Flowers (H&F)(note adapted to be stars and flowers for use in Africa)<br><br>Inhibit interference: MSIT<br><br>The flanker task requires (spatial) | The *PLUS-EF* tablet-based executive function tool, is an open-source android-based cognitive assessment tool that has been validated for school-aged children[16]. It has been adapted for use in the *PLUS -EF* tablet tests in urban Kenya[50]. It measures executive function including cognitive flexibility and inhibition using different tasks, of which 3 were used: Multi-source interference test, stars and flowers and flanker test. These tasks have been adapted for use in the *PLUS -EF* tablet tests in Kenya after extensive | Each individual test has shown reliability for an individual assessment situation, with quoted Cronbach alpha of MSIT 0.91 and H&F 0.81 [16].<br><br>A separate analysis of the Flanker task has shown good test-retest reliability and internal consistency, with Cronbach alpha >0.8[53]. | MSIT performance has been shown to improve with age and brain function mapped using functional MRI[54]. The MSIT has also previously demonstrated an effect of socioeconomic status, subjective social status and perceived stress on children's executive function[55].<br><br>Hearts and Flowers has previously shown detrimental effects of moving home and socioeconomic status[56] as well as positive impacts of schooling exposure with time[57]. |

[1] Note that the Plus EF tool was only added towards the end of this study and so data for Plus EF are not presented in this paper.

| Domain | Sub-domain | Tool | Outcome(s) | Validity | Reliability | Responsiveness |
|---|---|---|---|---|---|---|
| | | | selective attention and executive control. | piloting[50]. [Note this only got added later so data are not presented in this paper].<br><br>Similar tests have been widely used across childhood, although mainly in high-income settings[51]. The MSIT has been shown to reliably activate the cingulo-frontal-parietal (CFP) cognitive/attention network by functional MRI[52]. | | The Flanker test has previously showed improved selective attention with age in 4 to 6 year olds[58] and its inhibitory control is closely associated with school readiness[59]. |
| Cognitive Function | Fine motor | Finger tapping | Time to perform sequential finger tapping | *The Rapid Sequential Continuous Movements* was a sensitive measure of fine motor skills that was associated with stunting in children in Jamaica[24]. | During development, test-retest reliability was >0.78 and inter-observer agreements were above 0.96[60]. | An impact of stunting was shown, as well as strong associations with a schooling achievement test and intelligence quotient (IQ)[60]. |
| Cognitive Function | Academic function | School achievement test | Numeracy, literacy, Writing ability | Numeracy were assessed using elements from the Early Grade Maths Assessment[9] and UNICEF Multi-Indicator Cluster Survey (MICS) Foundational Learning Module[61], which had been widely applied across Zimbabwe.<br>Literacy was assessed using reading elements from *The Early Grade Reading Assessment (EGRA),* which has been widely used across Africa to assess literacy[9].<br>Name writing ability has been shown to associated with emergent literacy skills[10]. | EGMA report reliability with Spearman's rho of above 0.94 for number identification, discrimination and missing number subtests, with Cronbach alpha of 0.94, 0.82 and 0.58 respectively[13].<br>MICS used very similar questions with strong inter-rate reliability and agreement with EGRA and EGMA tests[62].<br>EGRA report reliability in Liberia of the 3 main elements used: letter identification, familiar word reading and non-word reading had Cronbach Alpha values of 0.78, 0.74 and 0.80 respectively[63]. | EGRA has been previously used to assess individual levels of literacy in Kenya[64].<br>EGRA has been used to monitor also early grade reading interventions[65].<br>The overall structure of the test, has been similarly piloted in Bangladesh and is being used to assess children followed up in the WASH Benefits trial [66] (Tofail, personal communication). |
| | Socioemotional function | Strengths and Difficulties Questionnaire | Total difficulties score | *The Strengths and Difficulties Questionnaire (SDQ)* is a brief screening caregiver questionnaire for child mental | Parent-reported total difficulties scores gave a Cronbach alpha of >0.77 in a large sample in Holland, | This tool recently demonstrated the impact of LNS in similar aged children in Ghana[69]. |

| Domain | Sub-domain | Tool | Outcome(s) | Validity | Reliability | Responsiveness |
|---|---|---|---|---|---|---|
| | | | | health and behavioural problems from age 3-16 years. Both parent and teacher version have been shown to have construct and prediction validity in Holland[67]. It has been widely used in Africa[23] and found to be highly acceptable and applicable in sub-Saharan African settings [68]. | although sub-scores were lower (0.42 to 0.8) [67]. | |
| Cognitive Function | Sensory and overall function | WG UNICEF child functioning module (CFM) | Overall score | This tool was developed across multiple countries by UNICEF with extensive pretesting, cognitive interviewing and adaption[70 71]. | The child functioning module was successfully performed in Mexico, Samoa and Serbia. It provided consistent prevalence rates similar to other tools using cut-offs describing 'a lot of difficulty' in functional domains or 'daily' levels of anxiety[72]. | The CFM has also been previously used in the SHINE cohort at age 2 years and demonstrated good agreement in comparison with functional screening using the Malawi Development Assessment Tool (MDAT) [73 74]. |
| Growth | Body composition | Bioimpedance | Impedance index (relative lean mass)<br>Lean mass index, Phase angle | *Bioimpedance (BIA)* measures tissue health and the proportion of lean mass using an imperceptible electrical signal between the hand and foot[75]. This has been widely used globally to assess malnutrition[76], and the technique has been validated and calibration equations derived using other body composition techniques such as deuterium dilution[77] in the Gambia. | | Bioimpedance has been used to show accretion of lean mass in children recovering from severe acute malnutrition (SAM)[76]. It has also been previously used in the SHINE study and showed a reduction with stunting (unpublished data).<br>Lean mass correlates with organ size [78], improved neurodevelopment [79] and reduced metabolic risk[80]. An Ethiopian birth cohort study showed an association between lean mass at birth and socio-emotional function measured using the SDQ[38]. |
| Growth | Body composition | Skinfold thickness | | *Skinfold thickness* measures the subcutaneous fat layer around the body and describes its distribution. Triceps and maximal calf skinfold thicknesses give a | Acceptable inter-observer agreement using technical error of measurement was <1mm, within the ChroSAM study[81]. | Fat mass provides short-term benefits for survival [82], but has longer-term metabolic health costs. Skinfold thickness and its distribution is a useful measure that |

| Domain | Sub-domain | Tool | Outcome(s) | Validity | Reliability | Responsiveness |
|---|---|---|---|---|---|---|
| | | | | measure of peripheral fat, whilst subscapular skinfolds measure central fat. | | reflects child malnutrition[76] and also as a marker for chronic disease risk[83]. They have also been previously used in the SHINE study and showed a reduction with stunting and HIV-exposure (unpublished data). |
| Growth | Anthropometry | | Height, Weight, Head circumference, Waist circumference, Hip circumference, Mid-upper arm circumference, Calf circumference, | *2d) Anthropometry*: Height and weight provide body mass index (BMI) which is an important marker of metabolic health[83], together with waist circumference[84]. Head circumference is a reliable measure of brain growth and previous nutritional deprivation, and is highly correlated with neurodevelopment[85]. Calf circumference and mid-upper arm circumference are complementary to skinfold thicknesses in providing insights into the quality of growth[86]. | Intra-observer technical error of measurement varied between 1 to 7 mm in the ChroSAM study[81]. | |
| Growth | Knee-heel length | | Knee-heel length | *Knee-heel (tibial) length* is a more sensitive measure of poor growth than leg length or stature and hence may be disproportionately reduced in stunting[87]. | | There is emerging evidence that knee-heel length is a proxy for organ size, e.g. kidney in stunted children [88]. It has also been previously used in the SHINE study and showed a reduction with stunting and HIV-exposure (unpublished data). |
| Physical function | Strength | Handgrip strength | Average Handgrip strength | Handgrip strength is one of the core tests both within the ALPHA[29] and PREFIT[31] test batteries. This has been selected from a systematic review of the literature,[32] combined with what had been previously piloted in Malawi (Kerac, personal communication). | Previous studies had shown high reliability coefficients=0.97 and 0.98 for right and left hands, respectively, and no difference between test and retest[29 89]. | *Handgrip strength* can be reduced in stunting[90] and in long-term assessments after acute malnutrition[86]. |
| Physical function | Strength | Broad jump | Distance jumped | Broad jump is one of the core tests both within the ALPHA[29] and PREFIT[31] test batteries. This was similarly | The broad jump has demonstrated good criterion validity and reliability. It had the strongest | *The broad jump* is a measure of truncal tone and fitness[90] and has been shown to |

| Domain | Sub-domain | Tool | Outcome(s) | Validity | Reliability | Responsiveness |
|---|---|---|---|---|---|---|
| | | | | recommended from systematic review of the literature,[32] as a validated test of core muscular fitness[91]. It had been previously piloted in Malawi (Kerac, personal communication). | association with a range of both lower body muscular strength tests (eg vertical jump, squat jump and countermovement jump) and upper body strength tests (throw basketball, push ups and isometric strength)[91]. | be reduced in stunted children in South Africa[92]. |
| Physical function | Cardiovascular fitness | Shuttle Run Test | Level reached | *The 20 meter shuttle run test (SRT)* is used to measure physical and aerobic capacity[93]. It has been shown to have good criterion related validity for cardiorespiratory fitness in both adults and children[94]. The criterion validity of the 20 m shuttle run test has been shown to be superior to similar measures of cardiovascular fitness such as the mile walk/run test[29 95]. | Reliability has been stated to be acceptable with no systematic bias[29 95]. | Stunting was a strong predictor of decreased fitness in the beep test when applied in Kenya[96] |
| Blood Pressure | | | | BP can be increased in stunting, particularly in combination with overweight[97]. | | BP in 8 year-olds in Nepal was independently negatively associated with leg and kidney length[78], and is a marker of homeostatic reserve and later cardio-metabolic risk[98]. |

Supplementary Table S1: Individual tools used within the SAHARAN toolbox, with outcomes, validity, reliability and responsiveness based on previous literature.

### Detailed methods

#### 1. Cognitive function

| | MARKER | MEASURE | Primary outcome | Secondary outcomes | RATIONALE |
|---|---|---|---|---|---|
| **Cognitive Function (120 mins)** | **K-ABC2** | Memory, spatial abilities, reasoning | Mental Processing Index (MPI) | Subtest scores | Measures cognitive processing: less schooling dependent |
| | **School Achievement Test** | Total score | *Subtest scores* | Academic | Literacy & numeracy |
| | **Fine motor** | Sequential finger tapping speed | Combined time to complete 2 tasks | Finger tapping | Fine motor |
| | **Plus-EF** | Executive Function | Accuracy in 3 executive tests | Individual subtest scores, reaction time | Executive function |
| | **Child socio-emotional questionnaire** | Home support | Total score | | Child's own perspective on home environment |
| | **Washington Group Child function module (asked in caregiver questionnaire)** | Disability screening, including vision and hearing | Overall score | Hearing, vision, mobility problem subscale | Child functional abilities |
| | **SDQ (asked in caregiver questionnaire)** | Socioemotional function | SDQ total score | SDQ subtest scores | Behaviour |

**Supplementary Table S2: cognitive measurement tools and outcomes used in the SAHARAN toolbox**

1.1 The 8 core subtests within the *Kaufmann Assessment Battery for Children 2nd Edition (K-ABC2)* were the primary outcome for cognition: Their scaled sum provided the mental processing index (MPI). It is available from www.pearson.com. The subtests selected were Atlantis, Story completion, Number recall, Atlantis delayed, Rover, Triangles, Word Order and pattern reasoning[43]. Online training was kindly provided by data collectors and trainers on zoom based in Uganda, Zimbabwe[48] and South Africa[43].

1.2 *The School Achievement Test* design was guided from piloting and similar tests currently being used in school-aged follow-up of the WASH Benefits trial[66] (Tofail F, private communication). During the test, the child started with numeracy, then moved to reading letters, then syllables and then words in the child's preferred language (79/80 children chose Shona, 1 chose English).

1.3 *Fine motor function was* guided by sharing training videos during pre-testing (Dr Chang-Lopez, private communication). The shortest time to complete the task of sequential finger tapping six times was the primary outcome. The data collector demonstrated first, then the child did several practice sessions before doing it 3 times in a row to ensure they could perform sequential finger tapping without stopping. Then each child was timed to see how fast they did sequential finger tapping 6 times in a row. They repeated this for a total of 3 times on each hand and the fastest time was used for analysis. The average between the 2 fastest times for each hand was also calculated.

1.4 The *PLUS-EF* tablet-based executive function tool, is an open-source android-based cognitive assessment tool. It measures executive function including cognitive flexibility and inhibition using 4 different tasks, of which 3 were used in this study (Multi-Source interference test (MSIT), hearts and stars, and flanker subtests) and their combined score was a primary outcome . Before each task, the child did training subtests with the data collector supporting with explanations and demonstrating how to hold the tablet in a standardised way (one hand each side of the tablet with thumbs free). The child did training subtests on the tablet which provided instant feedback to ensure standardised training and understanding before each task. The total Plus-EF score was calculated by summing the scores from all the tests taken together (excluding the practice tests). [Note that the Plus-EF was only added towards the end of this study, so data not shown].

1.5 *The Child Socioemotional Questionnaire* investigated the child's viewpoint on their socioemotional support within the home, together with one final question on food security. Four of the questions were previously used during an evaluation of a teacher's program in Zambia (MPES)[99]. In addition, 2 questions were previously used during a pilot study for UNICEF called Healthy Promoting Schools (HPS) (Dr Lisa Langhaug, private communication). All questions were cognitively interviewed and pre-tested before being used in the study.

1.6 The Washington Group / UNICEF child functioning module (WG) is an international screening tool used to identify children with disabilities[72]. This has been previously used in Zimbabwe and correlated with the validated Malawi Development Assessment Test (MDAT) score[74] at 24 months. It was used to assess caregiver-reported difficulties in hearing, vision, learning, communication or behaviour. A laminated pictorial Likert scale of answers was also used to help visualise responses.

*1.7 The Strengths and Difficulties Questionnaire (SDQ)* is a brief screening caregiver questionnaire for child mental health and behavioural problems from age 3-16 years. The SDQ asks the caregiver to describe their child's behaviour during the past 6 months, using 25 questions. Responses were scored on a Likert scale from 0-2 to give a "Total difficulties score" which was a primary outcome. The tool in English is free to download online (https://www.sdqinfo.com/). The total difficulties score was a primary outcome. A laminated pictorial Likert scale of answers was used to assist the caregiver with choosing a response.

#### 2. Body composition and anthropometry

| | MARKER | MEASURE | Primary outcome | Secondary outcomes | RATIONALE |
|---|---|---|---|---|---|
| **Body composition (20 mins)** | **BIA** | Impedance of tissues | Lean mass index, Phase angle | Impedance index | Quality of growth / metabolic health |
| | **Knee-heel length** | Tibial growth | Median Knee-heel | | Prioritization of growth |
| | **Triceps, scapular, calf skinfolds** | Subcutaneous fat | Sum of skinfolds | Individual skinfolds, Peripheral: central | Fat: peripheral c.f. central / metabolic health |

| | | | | | |
|---|---|---|---|---|---|
| **Anthropometry (15 mins)** | **Height, weight** | Growth | BMI | HAZ, WAZ, | Metabolic health |
| | **Head circ** | Brain volume | Head circumference | - | Prioritization of growth |
| | **Waist circ, Hip circumference** | Abdominal size | Waist circ | Hip circumference | Metabolic health |
| | **Calf circ, MUAC** | Peripheral fat / muscle | Calf circ, MUAC | - | Quality of growth |

**Supplementary Table S3: Tests of body composition and anthropometry in the SAHARAN toolbox**

*2a) Bioimpedance (BIA)* gives an impedance reading (*Z*), and $Height^2/Z$ gives the 'impedance index', which is a relative measure of lean mass within the sample. However, this requires a population-specific equation to provide the actual lean mass using another body composition technique[77]. The Lean mass index (1/Z) avoids the need for any equation and can be visualised as expressing variability in the lean mass component of body mass index, ie lean mass index expressed in the same $kg/m^2$ units. Note that 1/Z is expressed in abstract units (1/Ohms), but the variability in 1/Z and the variability in lean mass index differ only in terms of the constants used to convert abstract to physical values[100]. This makes 1/Z highly effective at ranking variability in lean mass index. BIA was measured in recumbent children using the Bodystat 1500 MDD instrument (BodyStat, Isle of Man, UK). Electrodes were attached to the right hand and foot and BIA measurements performed with standard criteria to ensure repeatability. The mean of the two readings was used for each BIA measurement. Data were excluded if the phase angle was more than 8 degrees[101], or Z had poor repeatability (> 6 Ohms difference). BIA assessments were tolerated extremely well by children in the study.

*2b) Skinfold thickness* was measured to the nearest 0.2mm using a skinfold caliper (Holtain, Crosswell, Wales) and the median of 3 readings used. These were similarly tolerated extremely well by children in the study. All measurements were performed on the right side.

*2c) Knee-heel (tibial) length*: The right-sided median tibial length was assessed using a commercial knemometer (weighandmeasures.com, Olney, USA).

*2d) Anthropometry*: Height was measured using a Shorrboard (Weighandmeasure, USA), weight using portable scales (Seca, Germany) and circumferences using anthropometry tape (Weighandmeasure, USA).

#### 3. Physical Function

| | MARKER | MEASURE | Primary outcome | Secondary outcomes | RATIONALE |
|---|---|---|---|---|---|
| **Physical Function (30 mins)** | **Grip strength** | Lean muscle both hand | Highest grip strength, average in both hands: a | Dominant and non-dominant hand strength | Lean muscle: hand |
| | **Broad jump** | Truncal muscles | Full distance: **b** | | Lean muscle: trunk |
| | **20m Beep test (composite score =PF)** | Physical Fitness, | Fitness level: **c** ***Composite score = a+b+c*** | Heart rate (HR) variability HR after 1 minute | Stamina, Overall composite score |
| | **Hemoglobin** | Anaemia | Hb | | Physical fitness |
| | **BP** | Fitness | Resting Systolic, diastolic bp | Pulse pressure, BP 1 minute after exercise | Cardiovascular fitness |

**Supplementary Table S4: Physical function measurements used in the SAHARAN toolbox**

*3a) Handgrip strength:* The Takei dynamometer was selected, as it was shown to have the highest criterion-related validity and reliability[102]. The dynamometer's handgrip size was adjusted appropriately for the handspan[89]. It has also been reported that use with the elbow extended provides the most appropriate positioning[102]. Therefore the child stood and held the Takei dynamometer vertically down and squeezed as hard as they could for up to 5 seconds. After a suitable break, they repeated this three times for each hand and the maximum value used for analysis.

*3b) The broad jump:* The child stands behind a line marked on the ground with feet slightly apart. A two-foot take-off and landing is used where the child swings the arms and bends the knees to provide forward drive, with the research nurse demonstrating first. The distance jumped is defined from the take-off line to the nearest point of contact on the landing (back of the heels), with the longest jump of 3 attempts used in analysis.

*3c) Shuttle run test (SRT):* A 20 meter tape measure is placed and the child runs repeatedly between each end, arriving before the beep, with increasingly shortened time gaps between beeps. A Bluetooth speaker is connected to the ODK tablet using the "Beep test" free android app (Beeptest, Ruval Enterprises, Canada). Once the child misses the beep three times in a row or stops running, the child then withdraws. The test provides a valid and reliable prediction of the $VO_{2max}$[103], the maximum rate at which the body uses oxygen during exercise. The feasibility of heart rate monitoring using wearable wrist (Fitbit Charge, UK) and chest-based (Polar, UK) heart rate monitors during exercise was also explored[104]. *A composite total fitness score* was calculated based on the standardised results of highest handgrip score, furthest broad jump and highest level on 20m beep test to give an overall measure of physical function.

*3d) Blood pressure* (BP) was initially measured using an automated sphygmomanometer (Omron, Milton Keynes, UK) and then manual blood sphygmomanometer (Medisave, UK).

*3e) Haemoglobin* was measured (Hemocue) by a finger prick test as a potentially important contributor to physical fitness[86] and cognitive outcomes.

#### 4. Caregiver questionnaire

In parallel with the child measurements, a detailed caregiver questionnaire was also administered to measure household demographics, previous adversities and contemporary factors associated with child growth and function.

| | MARKER | MEASURE | Primary outcome | Secondary outcomes | RATIONALE |
|---|---|---|---|---|---|
| **Caregiver questionnaire (90 mins)** | **Demographics** | Household composition | Main caregiver | Years of schooling | |
| | **Socioeconomic status** | SES score | Overall score | - | Socio-economic status |
| | **SDQ** | Socioemotional function | SDQ total score | SDQ subtest score | Behaviour |
| | **Schooling & COVID impact** | School engagement & attendance | Years of schooling | Attendance<br>Alternative learning | Education |
| | **Washington Group Child function module** | Disability screening, including vision and hearing | Overall score | Hearing, vision, mobility problem subscale | Child functional abilities |

| | | | | | |
|---|---|---|---|---|---|
| | **Child adversity scale** | Adversities | Overall score | | Measure of accumulated adversities |
| | **Child parent relationship scale** | Caregiver's relationship with child | Overall score | | Nurturing |
| | **MICS Child discipline score** | Caregiver's relationship with child | Overall score | | Nurturing |
| | **EPDS** | Maternal depression | Overall score | - | Depression |
| | **HFIAS, HDDS** | Food insecurity | HDDS score<br>HFIAS score | | Food insecurity |
| | **HWISE, Water access** | Water insecurity & access | HWISE score, water volume ad | | Water insecurity |

**Supplementary Table S5: Caregiver questionnaire sections. MICS: Multi-indicator cluster survey (UNICEF), EPDS: Edinburgh postnatal depression score, HFIAS: Household Food Insecurity Assessment Scale, HDDS: Household Dietary Diversity scale, Household Water Insecurity Experiences Scale (HWISE)**

*Demographics* related to household composition and the primary caregiver. *Socioeconomic status* was measured using a wealth index previously developed for the region of the study[105]. *Schooling exposure and COVID impact* were also asked. *The Washington Group / UNICEF Child Functioning module*[4 106] provides a screening tool for functional difficulties in hearing, vision, communication, comprehension, learning, mobility and emotions using a rating scale. *The child adversities index* screened for major life adversities associated with reduced child development since birth[107 108]. These questions were carefully piloted and selected for a region with minimal social services support. Key elements of nurturing were measured using the Child-Parent Relationship scale[109] and MICS Child Discipline questionnaire[61 110]. Caregiver depression was measured using local translations of The Edinburgh Postnatal Depression score (EPDS), which has been validated and extensively used in this region[111]. Food insecurity was measured using the Household Food Insecurity Assessment Scale (HFIAS)[112] and Household Dietary Diversity Scale (HDDS)[113]. The Household Water Insecurity Experiences Scale (HWISE)[114] measured water insecurity.

### Supplementary results tables

Additional associations within the SAHARAN toolbox were measured by least squares linear regression analysis as described below:

| Dependent variable | Independent variable | Regression Coefficient | Lower 95% CI | Upper 95% CI | P value |
|---|---|---|---|---|---|
| **Impedance Index** | **WAZ** | 0.23 | 0.19 | 0.28 | <0.001 |
| **HAZ** | **WAZ** | 0.67 | 0.52 | 0.83 | <0.001 |
| **Knee heel length, cm** | **WAZ** | 1.58 | 1.15 | 2.02 | <0.001 |
| **Head circumference, cm** | **WAZ** | 0.53 | 0.07 | 0.98 | 0.023 |
| **Phase angle, degrees** | **WAZ** | 0.26 | 0.11 | 0.40 | 0.001 |
| **Impedance Index** | **HAZ** | 0.20 | 0.14 | 0.25 | <0.001 |
| **Knee heel length, cm** | **HAZ** | 2.28 | 1.99 | 2.57 | <0.001 |
| **Head circumference, cm** | **HAZ** | 0.34 | -0.14 | 0.83 | 0.16 |
| **Phase angle, degrees** | **HAZ** | 0.04 | -0.12 | 0.20 | 0.61 |
| **Knee heel length, cm** | **Impedance Index** | 4.76 | 3.29 | 6.22 | <0.001 |
| **Impedance Index** | **Phase angle** | 0.12 | 0.03 | 0.22 | 0.01 |
| **LMI, Kg /m2** | **Phase angle** | 0.68 | 0.16 | 1.19 | 0.01 |
| **BMI Z-score** | **LMI** | 0.38 | 0.27 | 0.50 | <0.001 |
| **Impedance Index** | **Total skinfolds** | 0.01 | 0.00 | 0.02 | 0.10 |
| **Phase angle, degrees** | **Total skinfolds** | 0.02 | -0.01 | 0.05 | 0.29 |
| **BMI-Z** | **Total skinfolds** | 0.11 | 0.07 | 0.15 | <0.001 |

**Supplementary table S6**: Linear regression relationships between growth and body composition variables. Total sft: Total skinfold thickness, LMI: lean mass index

| Dependent variable | Independent variable | Coefficient | Lower 95% CI | Upper 95% CI | P value |
|---|---|---|---|---|---|
| **Maximum grip strength, Kg** | **HAZ** | 1.24 | 0.70 | 1.78 | <0.001 |
| **Maximum Broad jump, cm** | **HAZ** | 6.72 | 2.57 | 10.87 | <0.001 |
| **Shuttle run test level** | **HAZ** | 0.39 | 0.07 | 0.71 | 0.02 |
| **Maximum grip strength, Kg** | **WAZ** | 1.17 | 0.65 | 1.69 | <0.001 |
| **Maximum Broad jump, cm** | **WAZ** | 5.22 | 1.17 | 9.26 | 0.01 |
| **Shuttle run test level** | **WAZ** | 0.30 | -0.01 | 0.61 | 0.05 |
| **Total Physical Score** | **WAZ** | 1.08 | 0.55 | 1.61 | <0.001 |
| **Total Physical Score** | **MUAC, cm** | 0.44 | 0.07 | 0.82 | 0.02 |
| **Total Physical Score** | **Hip circumference, cm** | 0.18 | 0.06 | 0.30 | 0.004 |
| **Total Physical Score** | **Calf circumference, cm** | 0.34 | 0.08 | 0.60 | 0.01 |
| **Total Physical Score** | **Head circumference, cm** | 0.40 | 0.13 | 0.66 | 0.004 |
| **Total Physical Score** | **Impedance Index** | 4.50 | 2.91 | 6.08 | <0.001 |

| | | | | | |
|---|---|---|---|---|---|
| **Total Physical Score** | **Phase angle, degrees** | 1.14 | 0.33 | 1.95 | 0.006 |
| **Total Physical Score** | **Knee heel length, cm** | 0.49 | 0.29 | 0.70 | <0.001 |
| **Total Physical Score** | **Waist circumference, cm** | 0.13 | -0.03 | 0.29 | 0.12 |

**Supplementary Table S7: Relationships between growth and physical function measurements. MUAC: Mid-upper arm circumference, BMI: Body mass index**

| Dependent variable | Independent variable | Coefficient | Lower 95% CI | Upper 95% CI | P value |
|---|---|---|---|---|---|
| **MPI** | **SDQ** | -0.19 | -0.61 | 0.24 | 0.38 |
| **Fine motor** | **SDQ** | 0.16 | -0.15 | 0.47 | 0.30 |
| **SAT** | **Schooling years** | 18.71 | 13.99 | 23.44 | <0.001 |
| **MPI** | **Schooling years** | 2.65 | -0.11 | 5.41 | 0.06 |
| **Fine motor** | **Schooling years** | -0.90 | -3.21 | 1.42 | 0.44 |
| **SDQ** | **Schooling years** | -0.94 | -2.41 | 0.54 | 0.21 |
| **SAT** | **Child socioemotional** | 8.20 | 2.87 | 13.53 | 0.003 |
| **MPI** | **Child socioemotional** | 2.42 | -0.09 | 4.92 | 0.06 |
| **Fine motor** | **Child socioemotional** | -0.41 | -2.79 | 1.98 | 0.73 |
| **SDQ** | **Child socioemotional** | -0.12 | -1.49 | 1.25 | 0.87 |
| **SDQ** | **Total physical score** | -0.41 | -0.91 | 0.10 | 0.12 |
| **SAT** | **Total physical score** | 0.24 | -1.99 | 2.47 | 0.83 |

**Table S8: Linear regression relationships between cognition measurements, years of schooling, child socioemotional score and total physical score.**

### Supplementary figure

[please contact corresponding author for assistance with images]

**Fig S1. Application of the SAHARAN toolbox.** A: Portable handwashing station and role play. B: tent arrangement for caregiver and child (here using a tree for shade). C: Child cognitive measurement using the School Achievement Test (SAT). D: Child body composition measurement using Bioimpedance Impedance Analysis (BIA).
